# Is the United Kingdom (UK) medicines pricing policy failing patients? The impact of terminated National Institute for Health and Care Excellence (NICE) appraisals for multi-indication products on patients

**DOI:** 10.1101/2024.08.06.24311489

**Authors:** Helen Mitchell, Qian Xin, Jack Hide, Clement Halin, Swarali Sunil Tadwalkar, Sabera Hashim, Richard Hudson

**Affiliations:** Sanofi, Reading, UK; Clarivate, London, UK; University of Bristol, Bristol, UK

## Abstract

**Background:** National Institute for Health and Care Excellence (NICE) data regarding manufacturer-driven terminations indicate that some patients in the United Kingdom (UK) are unable to access treatments that are available in other European countries, which may result in reduced survival and quality of life (QoL). This study aims to quantify the health impact of NICE appraisals for multi-indication products terminated for reasons not related to clinical trial failure on the UK population.

**Methods:** Terminated NICE appraisals (2014–2023) for multi-indication products were identified and a targeted literature search was conducted to identify data on the health impact of the interventions. The potential incremental quality-adjusted life year (QALY) loss and impact on overall survival (OS), progression-free survival (PFS), and QoL was calculated.

**Results:** Over 16,000 QALYs/year were potentially lost (with one QALY equal to one year of life in perfect health) across approximately 829,000 patients in the UK due to NICE appraisals for multi-indication products being terminated for reasons not related to clinical trial failure. Across oncology indications (approximately 18,900 patients), OS and PFS may have been reduced by over 9,400 years and 9,000 years, respectively. The potential impact of the treatments for non-oncology indications for which NICE appraisals were terminated on QoL was an incremental improvement of 13% (weighted average).

**Conclusions:** Due to the increasing number of NICE terminations for multi-indication products, patients cannot access therapies that could lengthen their lives and increase their QoL. As the UK uniform pricing policy is likely to influence manufacturer-driven terminations, introducing alternative reimbursement arrangements such as indication-based pricing (IBP) agreements to ensure that prices remain commensurate with therapeutic value could improve access to therapies in the UK, thereby improving public health.

**Highlights:**

- National Institute for Health and Care Excellence (NICE) termination data indicate that some patients in the United Kingdom (UK) are unable to access treatments available in other European countries, which could potentially prolong their lives and improve their quality of life (QoL)
- Across approximately 829,000 patients in the UK, over 16,000 quality-adjusted life years (QALYs) per year (with one QALY equal to one year of life in perfect health) are potentially lost as a result of NICE appraisals for multi-indication products that have been terminated for reasons not related to clinical trial failure
- Assessing reimbursement options such as indication-based pricing (IBP) agreements for treatments that would typically not meet NICE’s cost-effectiveness criteria at the current price provides an opportunity to improve access to therapies in the UK, thereby improving public health

## Introduction

The National Institute for Health and Care Excellence (NICE), the health technology appraisal (HTA) body for England and Wales, technology appraisal process relies on companies submitting evidence, in line with NICE’s specification. If companies do not make a submission, which may be because the company feels that they cannot succeed with the appraisal, or if NICE is not satisfied that the evidence submission is adequate to reach a decision, then the appraisal is terminated. Consequently, NICE is unable to make a recommendation about the use of the technology in the National Health Service (NHS) and the medicine is not made routinely available (1). There has recently been a substantial rise in terminated NICE appraisals, with 16.6% of all appraisals for multi-indication and single-indication products terminated in 2017, increasing to 25.6% in 2022 (2). Terminations disproportionately impacted products with multiple indications; appraisals for multi-indication products accounted for 57.0% of all NICE submissions made post July 2016 to September 2023, but made up 63.9% of terminations (2). In 2022, appraisals for multi-indication products (n=37) accounted for 22% more submissions than single-indication appraisals (n=45), but resulted in double the number of terminations (2). Although the overall number of NICE terminations in 2023 fell slightly compared to 2022 (3, 4), the data still pose serious concerns about potential systemic pricing barriers affecting patient access in England and Wales. While some of these terminations may have been due to clinical trial failure, the rise in terminations indicates that companies may be increasingly choosing not to submit to NICE on the basis that demonstrating cost-effectiveness at the current price will be too difficult under the current system. The United Kingdom (UK) uniform pricing policy currently limits recognition of the therapeutic value of multi-indication products because the lowest cost-effective price determined for any indication must be implemented across all indications, including those already assessed by NICE (5). Consequently, the price of multi-indication products may not remain commensurate with therapeutic value. In practice this means that follow-on indications would have to be launched at loss despite increased numbers of patients receiving the drug overall. To prevent this, indication-based pricing (IBP) agreements could allow price to vary by indication, or take the form of a single price based on a weighted average of value and usage across indications, allowing the therapeutic value of each indication to be fully recognised (6). The issue has been highlighted in the recently published Commercial Framework for New Medicines (7), and grows ever more urgent as the number of treatments receiving regulatory approval for multiple indications increases; for example, 75% of all targeted treatments in oncology were licenced for multiple indications as of 2018 (8).

The increase in NICE terminations means that patients in the UK are missing out on innovative treatments that could improve their survival and increase their quality of life (QoL), and contributes to disparity with other European countries. For every 100 patients who access a new medicine in its first year of launch in other parts of the European Union (EU), just 21 patients in the UK receive access (9). In some cases, NICE terminations, which apply in England and Wales, have also created inequalities within the UK. Manufacturers are obliged to make a separate submission to the HTA body in Scotland, the Scottish Medicines Consortium (SMC), which has issued a positive opinion for a small number of medicines with no NICE recommendation. For example, tisagenlecleucel was accepted for use within NHS Scotland for the treatment of adult patients with relapsed or refractory diffuse large B-cell lymphoma (DLBCL) after two or more lines of systemic therapy under the end of life and ultra-orphan medicine process (2019) (10), but the corresponding NICE appraisal was terminated (2023) (11).

The standard measure used by NICE to determine the incremental benefit of one treatment over another is the quality-adjusted life year (QALY), where one QALY is equal to one year of life lived in perfect health. This provides a way of standardising assessments to ensure all treatments are judged fairly against a common unit. In the current study, the health impact of terminated NICE appraisals for products with multiple indications across multiple therapy areas was determined by calculating total annual QALYs foregone to the UK population, along with the impact on survival and QoL.

## Methods

NICE appraisals for products with multiple indications terminated over the past 10 years (2014–2023) for reasons not related to clinical trial failure were identified. As no manufacturer submissions providing health outcomes are available for terminated appraisals, a targeted literature search was conducted to identify data on the health impact of the interventions. The potential incremental QALY loss vs. standard of care (SoC) resulting from NICE terminations was estimated for all indications and, where data were available, additional qualitative analysis was conducted to explore other potential health impacts that could not be quantified at the population level due to a lack of data. The potential impact on overall survival (OS) and progression-free survival (PFS) was calculated for oncology indications, and the overall QoL impact was calculated for non-oncology indications.

### Identification of Terminated NICE Appraisals and Targeted Literature Searching

A “step-by-step” methodology was used to evaluate terminated NICE appraisals for products with multiple indications. The NICE website was searched to identify all terminated appraisals for multi-indication products over the past 10 years (2014–2023), excluding appraisals with evidence that termination was due to clinical trial failures, or appraisals followed by a resubmission.

A targeted literature search was conducted in February 2024 using PubMed and Google Scholar, to identify data on the health impact of the interventions belonging to the identified terminated appraisals. The eligibility criteria for publications related to the terminated indications are described in Table 1.

**Table 1:** Publication eligibility criteria.

| Category | Eligibility criteria |
| --- | --- |
| **Population** | For each indication, publications reporting outcomes on a UK population were prioritised where available:<br>• If data were not available for the UK population for a specific indication, publications from other (similar) countries reporting outcomes for the indication of interest were included<br>• If data were not available for the indication of interest, UK-based publications providing proxy data for other (similar) indications were prioritised<br>• If neither data for the specific indication of interest or the UK were available, data were sourced for similar indications and similar locations |
| **Study design** | CEAs/CUAs were prioritised. If no CEA/CUA was identified, literature reviews, individual RCTs, and individual RWE papers were considered for inclusion |
| **Outcomes** | Outcomes for each of the products in the indications for which their NICE appraisal was terminated:<br>• QALYs<br>• OS<br>• PFS<br>• QoL<br>• Other |

Abbreviations: CEA, cost-effectiveness analysis; CUA, cost-utility analysis; NICE, National Institute for Health and Care Excellence; OS, overall survival; PFS, progression-free survival; QALY, quality-adjusted life year; QoL, quality of life; RCT, randomised controlled trial; RWE, real-world evidence; UK, United Kingdom.

Where available, epidemiology data for the patient populations of interest in the UK were sourced from a proprietary database (12), and supplementary targeted literature searching was used to fill any gaps in the available data. For most interventions for which NICE appraisals were terminated, the corresponding SMC submission was also terminated or there was no submission to the SMC. Therefore, all population size estimates in this study were based on the UK perspective.

### Data Analysis

A proprietary database was used to determine the relevant drug-treatable incident/prevalent populations for all populations of interest, corresponding to the description in the terminated NICE appraisal (12). For all oncology indications, diagnosed incident populations, or populations based on relevant line of therapy, resectability status or stage of diagnosis, as described in the terminated appraisal, were estimated. For non-oncology indications, diagnosed prevalent populations, or corresponding populations with additional stratifications or a mutation positive treatable population, were estimated. United Nations projected population estimates were used to estimate the case counts for the year 2023 (13).

To determine the potential incremental QALY loss vs. current standard of care resulting from the identified NICE terminations, annual incremental QALYs were estimated for each indication using the total incremental QALYs and the time horizon reported in the literature for the indication. Where no time horizon was reported, a lifetime horizon was assumed. The actual time horizon (in years) for a lifetime model was estimated using both the national population life expectancy and the average age in the trial. Potential QALY losses were analysed both overall and within the oncology and non-oncology subgroups. To determine the potential impact on OS and PFS, mean survival was collected or estimated using published survival analyses for each oncology indication, and survival impact was aggregated to the population level based on population size. In cases where only median survival or survival rates at specific time points were provided, a simple exponential distribution was assumed to convert these values to mean survival, to facilitate analysis on a consistent scale. The UK general population life expectancy was included as an upper limit for the estimated PFS and OS in the analysis (14).

For the QoL analysis, considering that published studies used various disease specific QoL questionnaires, the absolute change in QoL scores is not informative. Therefore, the incremental percentage change in QoL scores from baseline was estimated. The overall QoL impact was calculated as the weighted average improvement, with weights assigned based on the estimated patient numbers for each indication. Additional qualitative analysis was conducted for all indications, where available, to explore other potential health impacts that could not be quantified at the population level due to a lack of data.

### Role of Funding

This study was funded by Sanofi, with funding used to conduct the analysis and for writing support.

## Results

### Terminated NICE Appraisals for Products with Multiple Indications

In total, 25 NICE appraisals for products with multiple indications meeting the inclusion criteria were terminated between 2014 and 2023 (Figure 1), 23 of which were included in the final analysis once duplicated indications were removed to avoid double counting (Table 2). The exclusion of duplicated indications was based on the availability, relevance, and quality of the collected evidence. Of these 23 appraisals, seven were in oncology indications and 16 were in non-oncology indications.

**Figure 1:**
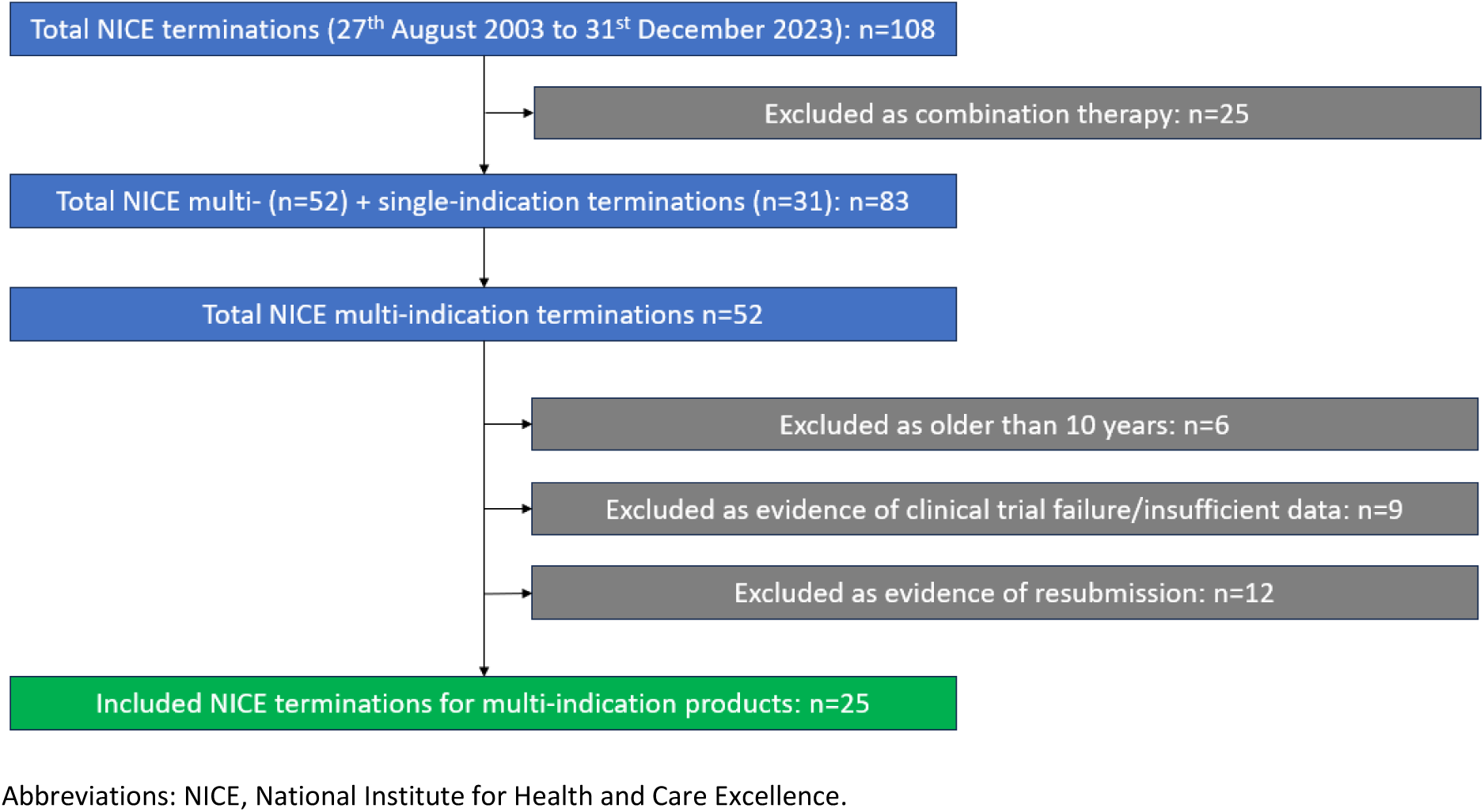
Selection of terminated NICE appraisals for products with multiple indications

**Table 2:**
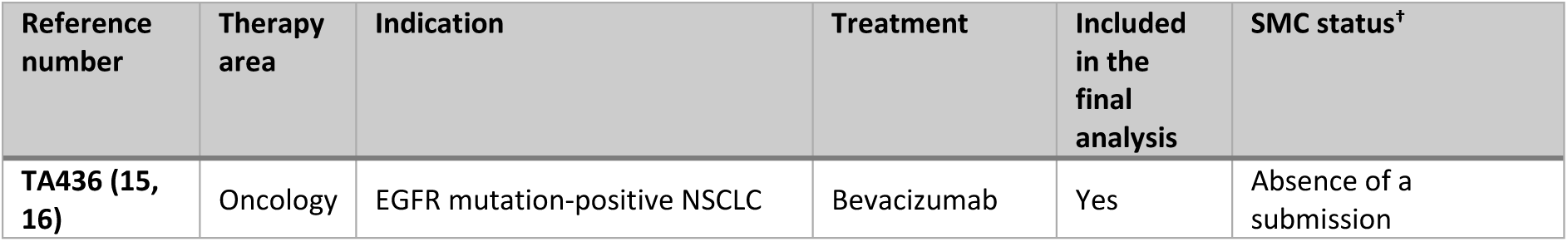
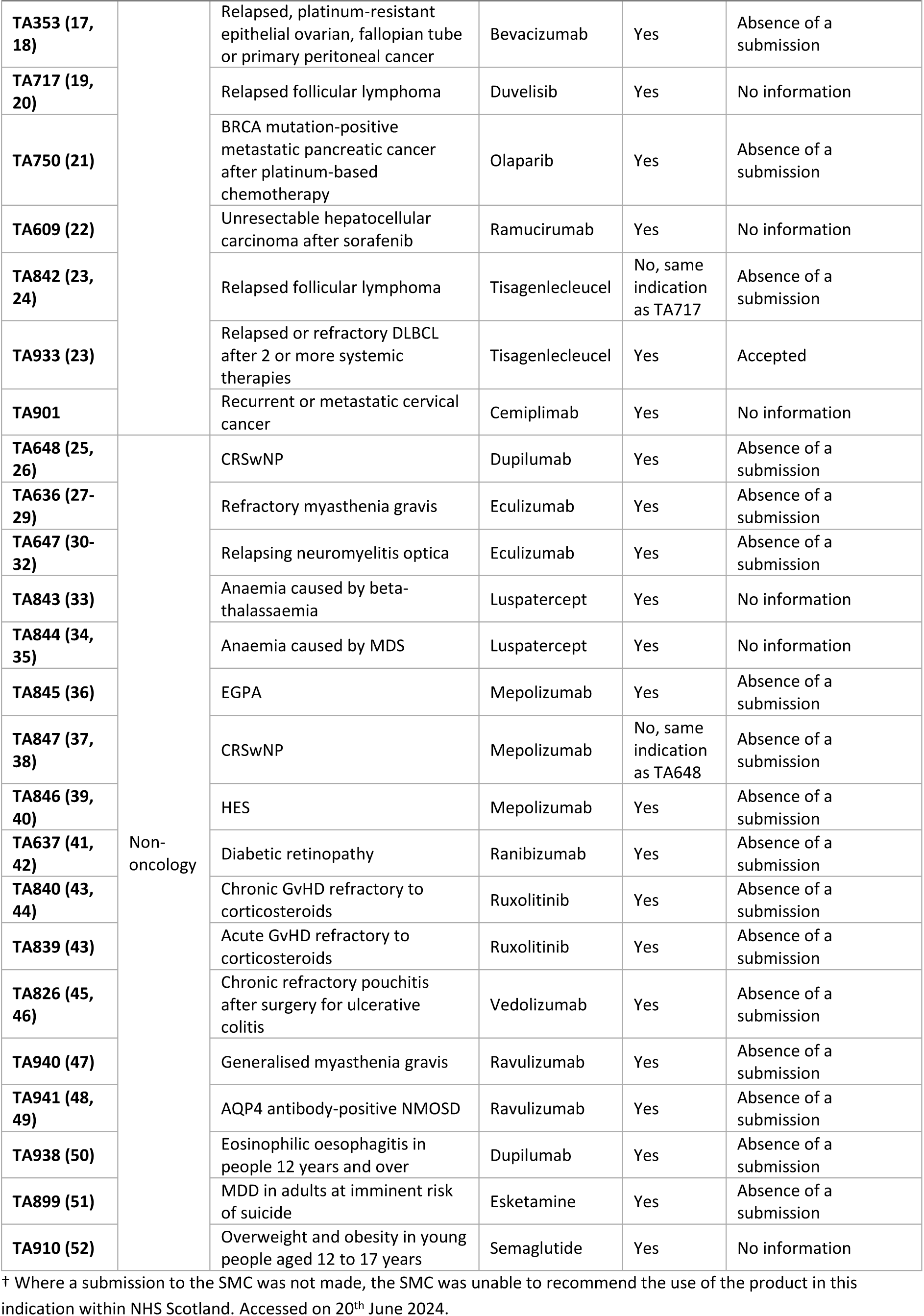
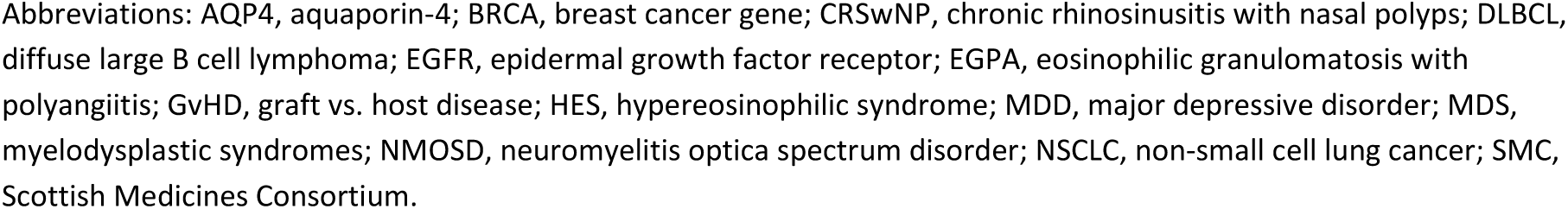
Summary of included NICE appraisals for multi-indication products.

| Reference number | Therapy area | Indication | Treatment | Included in the final analysis | SMC status<sup>†</sup> |
| --- | --- | --- | --- | --- | --- |
| TA436 (15, 16) | Oncology | EGFR mutation-positive NSCLC | Bevacizumab | Yes | Absence of a submission |
| TA353 (17, 18) |  | Relapsed, platinum-resistant epithelial ovarian, fallopian tube or primary peritoneal cancer | Bevacizumab | Yes | Absence of a submission |
| TA717 (19, 20) |  | Relapsed follicular lymphoma | Duvelisib | Yes | No information |
| TA750 (21) |  | BRCA mutation-positive metastatic pancreatic cancer after platinum-based chemotherapy | Olaparib | Yes | Absence of a submission |
| TA609 (22) |  | Unresectable hepatocellular carcinoma after sorafenib | Ramucirumab | Yes | No information |
| TA842 (23, 24) |  | Relapsed follicular lymphoma | Tisagenlecleucel | No, same indication as TA717 | Absence of a submission |
| TA933 (23) |  | Relapsed or refractory DLBCL after 2 or more systemic therapies | Tisagenlecleucel | Yes | Accepted |
| TA901 |  | Recurrent or metastatic cervical cancer | Cemiplimab | Yes | No information |
| TA648 (25, 26) | Non-oncology | CRSwNP | Dupilumab | Yes | Absence of a submission |
| TA636 (27-29) |  | Refractory myasthenia gravis | Eculizumab | Yes | Absence of a submission |
| TA647 (30-32) |  | Relapsing neuromyelitis optica | Eculizumab | Yes | Absence of a submission |
| TA843 (33) |  | Anaemia caused by beta-thalassaemia | Luspatercept | Yes | No information |
| TA844 (34, 35) |  | Anaemia caused by MDS | Luspatercept | Yes | No information |
| TA845 (36) |  | EGPA | Mepolizumab | Yes | Absence of a submission |
| TA847 (37, 38) |  | CRSwNP | Mepolizumab | No, same indication as TA648 | Absence of a submission |
| TA846 (39, 40) |  | HES | Mepolizumab | Yes | Absence of a submission |
| TA637 (41, 42) |  | Diabetic retinopathy | Ranibizumab | Yes | Absence of a submission |
| TA840 (43, 44) |  | Chronic GvHD refractory to corticosteroids | Ruxolitinib | Yes | Absence of a submission |
| TA839 (43) |  | Acute GvHD refractory to corticosteroids | Ruxolitinib | Yes | Absence of a submission |
| TA826 (45, 46) |  | Chronic refractory pouchitis after surgery for ulcerative colitis | Vedolizumab | Yes | Absence of a submission |
| TA940 (47) |  | Generalised myasthenia gravis | Ravulizumab | Yes | Absence of a submission |
| TA941 (48, 49) |  | AQP4 antibody-positive NMOSD | Ravulizumab | Yes | Absence of a submission |
| TA938 (50) |  | Eosinophilic oesophagitis in people 12 years and over | Dupilumab | Yes | Absence of a submission |
| TA899 (51) |  | MDD in adults at imminent risk of suicide | Esketamine | Yes | Absence of a submission |
| TA910 (52) |  | Overweight and obesity in young people aged 12 to 17 years | Semaglutide | Yes | No information |

<sup>†</sup> Where a submission to the SMC was not made, the SMC was unable to recommend the use of the product in this indication within NHS Scotland. Accessed on 20<sup>th</sup> June 2024.
Abbreviations: AQP4, aquaporin-4; BRCA, breast cancer gene; CRSwNP, chronic rhinosinusitis with nasal polyps; DLBCL, diffuse large B cell lymphoma; EGFR, epidermal growth factor receptor; EGPA, eosinophilic granulomatosis with polyangiitis; GvHD, graft vs. host disease; HES, hypereosinophilic syndrome; MDD, major depressive disorder; MDS, myelodysplastic syndromes; NMOSD, neuromyelitis optica spectrum disorder; NSCLC, non-small cell lung cancer; SMC, Scottish Medicines Consortium.

### Targeted Literature Searching

Thirty-six studies were included in the final analyses, including 19 cost-effectiveness analyses, six meta-analyses or literature reviews, six randomized control trials, two Institute for Clinical and Economic Review reports, one NICE technical appraisal, one indirect treatment comparison study, and one observational study. Most of the included studies were based in the UK or had an international scope, with a selection originating from the US, Canada, Japan, Singapore, and China. The full details of the included studies can be found in Appendix 1.

### Health Impact

#### Patient Demographics

Patient demographics for each analysis, aggregated across indications, are described in Table 3. The majority of patients included in the QALY analysis and the survival analysis (OS and PFS) were female, and the majority of patients included in the QoL analysis were male (Table 3). The mean age of patients included in the QALY analysis, the survival analysis, and the QoL analysis was 52 years, 66 years, and 50 years, respectively.

**Table 3:** Patient demographic information across included studies.

|  | Studies included in QALY analysis<sup>†</sup> | Studies included in survival analysis<sup>‡</sup> | Studies included in QoL analysis<sup>§</sup> |
| --- | --- | --- | --- |
| **Number of patients** | 5,237 | 1,851 | 4,775 |
| **Mean age, years<sup>¶</sup>** | 52 | 66 | 50 |
| **Female patients, %<sup>¶</sup>** | 60% | 68% | 27% |

<sup>†</sup> Patient demographics aggregated across all indications.
<sup>‡</sup> Patient demographics aggregated across all oncology indications.
<sup>§</sup> Patient demographics aggregated across all non-oncology indications.
<sup>¶</sup> Weighted average of all studies included in the analysis.
Abbreviations: QALY, quality-adjusted life year; QoL, quality of life.

#### Population Estimates

Relevant incident/prevalent populations for all populations of interest are presented in Table 4. As most treatments with terminated NICE appraisals for multi-indication products either had a corresponding terminated SMC appraisal or were not submitted to the SMC, all population size estimates were based on the UK perspective. The estimated total population of interest was 831,871 patients (n=18,887 for oncology, n=812,984 for non-oncology).

**Table 4:**
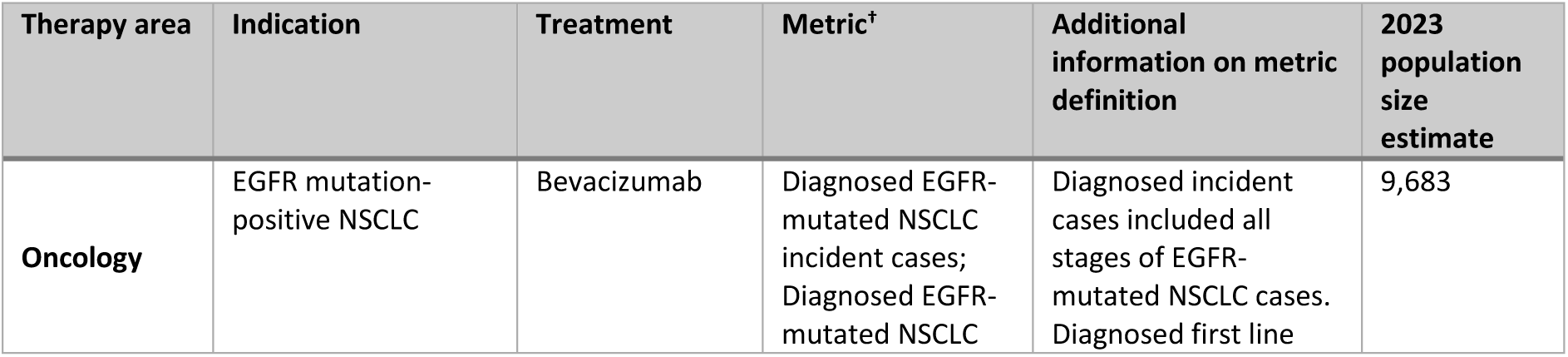
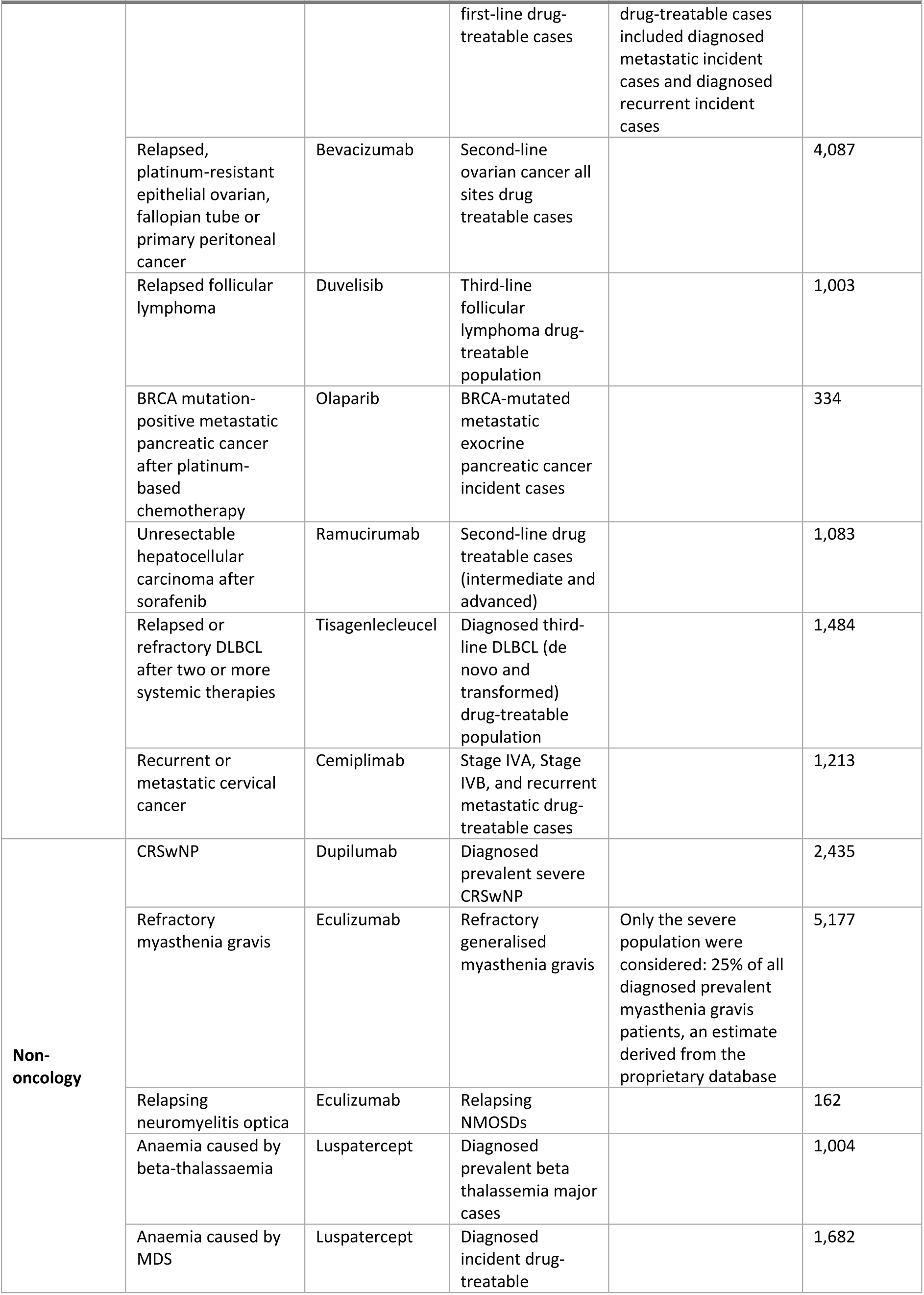
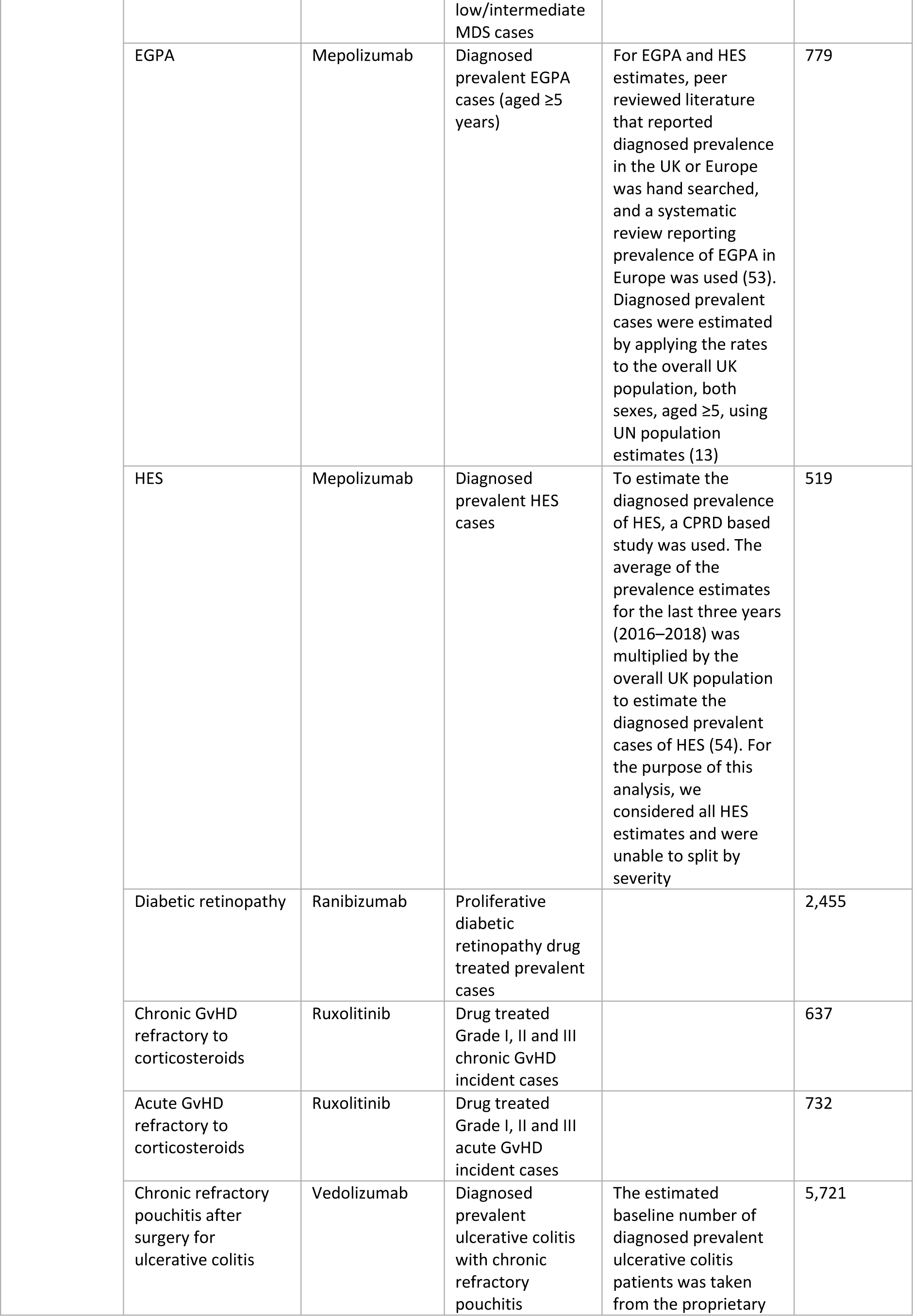
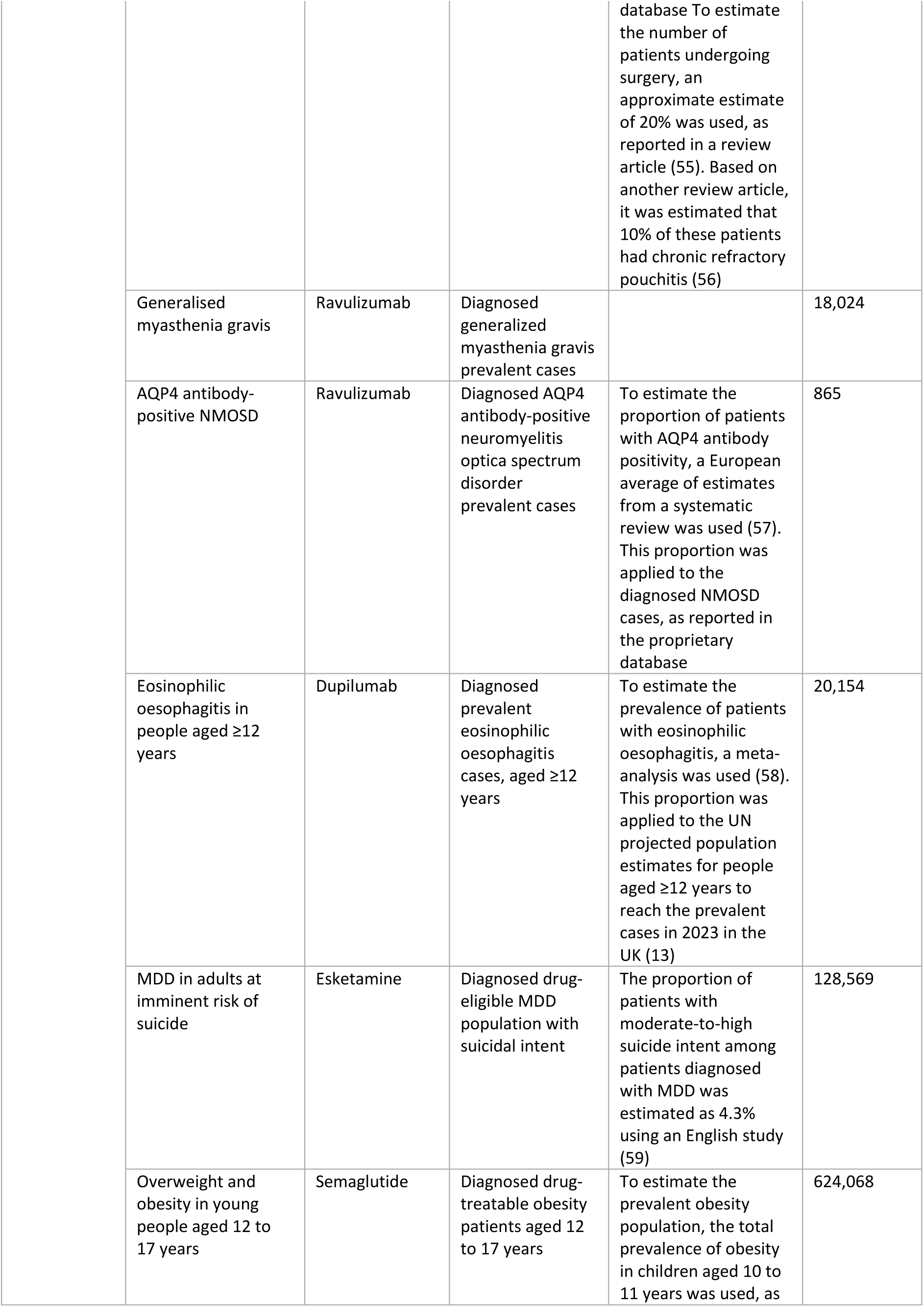
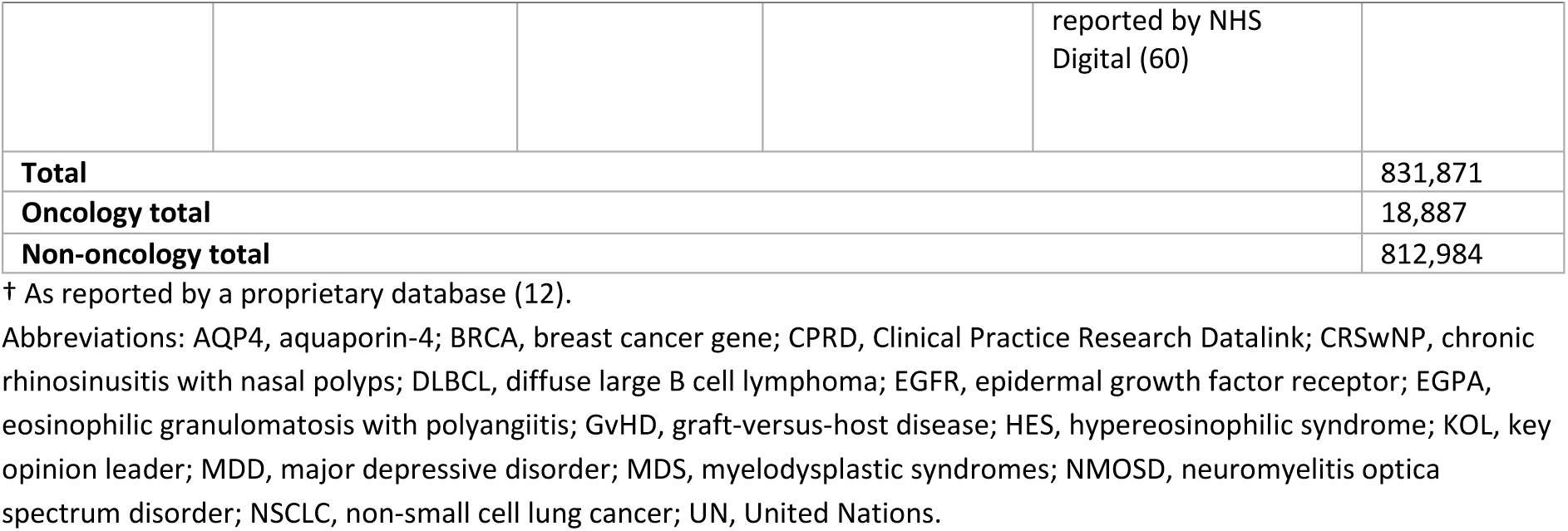
Population size estimates.

#### Impact on QALY

The potential incremental QALY loss resulting from the termination of NICE appraisals for multi-indication products for reasons not related to clinical trial failure was 16,079 QALYs/year, with 277 QALYs/year potentially lost for oncology populations and 15,802 QALYs/year for non-oncology populations (Table 5). The largest loss of incremental QALYs because of NICE terminations for any oncology indication was in the relapsed follicular lymphoma population, where an estimated 150 QALYs/year were lost. The largest loss of incremental QALYs because of NICE terminations for any non-oncology indication was in the overweight and obesity in young people aged 12 to 17 years population, where an estimated 9,361 QALYs/year were lost.

**Table 5:** Potential annual QALY loss.

| Therapy area | Indication | Annual discounted incremental QALYs<sup>†</sup> | Total annual population number | Population annual discounted incremental QALYs<sup>‡</sup> |
| --- | --- | --- | --- | --- |
| **Oncology** | EGFR mutation-positive NSCLC | -0.01 | 9,683 | -114 |
|  | Relapsed, platinum-resistant epithelial ovarian, fallopian tube or primary peritoneal cancer | 0.01 | 4,087 | 61 |
|  | Relapsed follicular lymphoma | 0.15 | 1,003 | 150 |
|  | BRCA mutation-positive metastatic pancreatic cancer after platinum-based chemotherapy | 0.07 | 334 | 23 |
|  | Unresectable hepatocellular carcinoma after sorafenib | 0.01 | 1,083 | 8 |
|  | Relapsed or refractory DLBCL after 2 or more systemic therapies | 0.08 | 1,484 | 113 |
|  | Recurrent or metastatic cervical cancer | 0.03 | 1,213 | 36 |
| **Non-oncology** | CRSwNP | 0.11 | 2,435 | 263 |
|  | Refractory myasthenia gravis | 0.08 | 5,177 | 388 |
|  | Relapsing neuromyelitis optica | 0.12 | 162 | 20 |
|  | Anaemia caused by beta-thalassaemia | NR | NR | NR |
|  | Anaemia caused by MDS | NR | NR | NR |
|  | EGPA | 0.05 | 779 | 42 |
|  | HES | 0.05 | 519 | 28 |
|  | Diabetic retinopathy | 0.01 | 2,455 | 17 |
|  | Chronic GvHD refractory to corticosteroids | 0.02 | 637 | 13 |
|  | Acute GvHD refractory to corticosteroids | 0.01 | 732 | 7 |
|  | Chronic refractory pouchitis after surgery for ulcerative colitis | 0.00 | 5,721 | 22 |
|  | Generalised myasthenia gravis | 0.08 | 18,024 | 1,352 |
|  | AQP4 antibody-positive NMOSD | 0.12 | 865 | 108 |
|  | Eosinophilic oesophagitis in people 12 years and over<sup>§</sup> | 0.12 | 20,154 | 2,380 |
|  | MDD in adults at imminent risk of suicide | 0.01 | 128,569 | 1,800 |
|  | Overweight and obesity in young people aged 12 to 17 years | 0.02 | 624,068 | 9,361 |
| **Total** |  | 1.14 | 829,185 | 16,079 |
| **Oncology** |  | 0.33 | 18,887 | 277 |
| **Non-oncology** |  | 0.80 | 810,298 | 15,802 |

<sup>†</sup> Rounded to two decimal places.
<sup>‡</sup> Rounded to zero decimal places.
<sup>§</sup> Sanofi, data on file (61).
Negative numbers indicate that a new drug was not as efficient as the current standard of care.
Abbreviations: AQP4, aquaporin-4; BRCA, breast cancer gene; CRSwNP, chronic rhinosinusitis with nasal polyps; DLBCL, diffuse large B cell lymphoma; EGFR, epidermal growth factor receptor; EGPA, eosinophilic granulomatosis with polyangiitis; GvHD, graft-versus-host disease; HES, hypereosinophilic syndrome; MDD, major depressive disorder; NMOSD, neuromyelitis optica spectrum disorder; NR, not reported; NSCLC, non-small cell lung cancer; QALY, quality-adjusted life year.

#### Impact on Survival

To determine the potential impact of NICE terminations on OS and PFS in oncology, mean survival was collected or estimated using survival analysis for each indication, and survival impact was aggregated to a population level based on population size. As a result of NICE terminations for multi-indication products due to reasons not related to clinical trial failure, OS and PFS in the overall oncology population were potentially reduced by 113,109 months (9,426 years) and 109,064 months (9,089 years), respectively (Table 6). The largest impact on OS was in the relapsed, platinum-resistant epithelial ovarian, fallopian tube or primary peritoneal cancer population, where OS was reduced by 48,324 months (4,027 years), and the largest impact on PFS was in the EGFR mutation-positive NSCLC population, where PFS was reduced by 77,819 months (6,485 years).

**Table 6:** Potential impact on OS and PFS (oncology only)

| Indication | PFS, mean<sup>†</sup> | OS, mean<sup>†</sup> | Total population, n | Population PFS, months<sup>‡</sup> | Population OS, months<sup>‡</sup> |
| --- | --- | --- | --- | --- | --- |
| EGFR mutation-positive NSCLC | 8.0 | 3.8 | 9,683 | 77,819 | 36,698 |
| Relapsed, platinum-resistant epithelial ovarian, fallopian tube or primary peritoneal cancer | 7.3 | 11.8 | 4,087 | 29,981 | 48,324 |
| Relapsed follicular lymphoma | -2.2 | 12.4 | 1,003 | -2,171 | 12,444 |
| BRCA mutation-positive metastatic pancreatic cancer after platinum-based chemotherapy | 5.2 | 1.2 | 334 | 1,735 | 385 |
| Unresectable hepatocellular carcinoma after sorafenib | 1.7 | 1.7 | 1,083 | 1,875 | 1,875 |
| Relapsed or refractory DLBCL after 2 or more systemic therapies | NR | 4.9 | 1,484 | NR | 7,258 |
| Recurrent or metastatic cervical cancer | -0.1 | 5.0 | 1,213 | -175 | 6,125 |
| **Total** | **20.0** | **40.8** | **18,887** | **109,064** | **113,109** |

<sup>†</sup> Rounded to one decimal place.
<sup>‡</sup> Rounded to zero decimal places.
Negative numbers indicate that a new drug was not as efficient as the current standard of care.
Abbreviations: BRCA, breast cancer gene; DLBCL, diffuse large B cell lymphoma; EGFR, epidermal growth factor receptor; NSCLC, non-small cell lung cancer; OS, overall survival; PFS, progression-free survival; NR, not reported.

#### Impact on QoL

The potential impact of the treatments for non-oncology indications for which NICE appraisals for multi-indication products were terminated for reasons not related to clinical trial failure on QoL was an incremental improvement of 13% (weighted average) (Table 7).

**Table 7:** Potential impact of the treatments for which NICE appraisals for multi-indication products were terminated for reasons not related to clinical trial failure on patient QoL.

| Indication | QoL Measure | QoL (incremental) |  | Estimated population number |
| --- | --- | --- | --- | --- |
|  |  | Incremental difference<sup>†</sup> | Incremental %<sup>‡</sup> |  |
| CRSwNP | SNOT-22 | 19.91 | 40% | 2,435 |
| Refractory myasthenia gravis | MG-QOL15r | 1.70 | 19% | 5,177 |
| Relapsing neuromyelitis optica | mRS | 0.33 | 17% | 162 |
| Anaemia caused by beta-thalassaemia | EORTC QLQ-C30 | 3.76 | 6% | 1,004 |
| Anaemia caused by MDS | EORTC QLQ-C30 | 3.76 | 6% | 1,682 |
| EGPA | NA | NA | NA | NA |
| HES | SF-36 | 0.04 | 4% | 519 |
| Diabetic retinopathy | VFQ-25 | 0.06 | 6% | 2,455 |
| Chronic GvHD refractory to corticosteroids | NA | NA | NA | NA |
| Acute GvHD refractory to corticosteroids | NA | NA | NA | NA |
| Chronic refractory pouchitis after surgery for ulcerative colitis | EQ-5D VAS | 0.13 | 13% | 5,721 |
| Generalized myasthenia gravis | MG-QOL15r | 1.70 | 19% | 18,024 |
| AQP4 antibody-positive NMOSD | mRS | 0.33 | 17% | 865 |
| Eosinophilic oesophagitis in people 12 years and over | NA | NA | NA | NA |
| MDD in adults at imminent risk of suicide | EQ-5D VAS | 4.70 | 11% | 128,569 |
| Overweight and obesity in young people aged 12 to 17 years | NA | NA | NA | NA |
| **Weighted average incremental %** |  | 13% |  |  |

<sup>†</sup> Rounded to two decimal places.
<sup>‡</sup> Rounded to zero decimal places.
Abbreviations: AQP4, aquaporin-4; CRSwNP, chronic rhinosinusitis with nasal polyps; EGPA, eosinophilic granulomatosis with polyangiitis; EORTC QLQ-C30, European Organization for the Research and Treatment of Cancer Core Quality of Life Questionnaire; EQ-5D VAS, EuroQol-5 Dimension Visual Analogue Scale; GvHD, graft-versus-host disease; HES, hypereosinophilic syndrome; MDD, major depressive disorder; MDS, myelodysplastic syndromes; mRS, Modified Rankin Scale; MG-QOL15r, Myasthenia Gravis Quality of Life-15 Item Scale revised; NMOSD, neuromyelitis optica spectrum disorder; NA, not available; QoL, quality of life; SF-36, Short-Form 36 Health Survey Questionnaire; SNOT-22, 22-item Sinonasal Outcome Test; VFQ-25, 25-Item Visual Function Questionnaire.

#### Impact on Other Health Outcomes

Additional qualitative analysis was conducted to explore other potential health impacts that could not be quantified at the population level due to a lack of data. This analysis indicated that the technologies from the terminated NICE appraisals for multi-indication products assessed in this study could potentially reduce the need for rescue therapy, surgery, hospitalization, and transfusions (29, 33, 34, 38). Additionally, some of these technologies have been associated with a minor positive effect on caregiver QoL and their ability to achieve major life goals related to education, work, or family life (27).

## Discussion

This analysis highlighted the impact of terminated NICE appraisals for multi-indication products on patient QALYs, survival, and QoL in the UK. The potential incremental QALY loss in the UK as a result of NICE appraisals for multi-indication products terminated for reasons not related to clinical trial failure was 16,079 QALYs/year across approximately 829,000 patients in the UK, with 277 QALYs/year potentially lost for oncology populations (approximately 18,900 patients) and 15,802 QALYs/year for non-oncology populations (approximately 810,000 patients). As a result of terminated NICE appraisals for multi-indication products for oncology indications, the mean OS and PFS in the overall oncology population were potentially reduced by 113,109 months (9,426 years) and 109,064 months (9,089 years), respectively, with the largest impact on OS in the relapsed, platinum-resistant epithelial ovarian, fallopian tube or primary peritoneal cancer population and the largest impact on PFS in the EGFR mutation-positive NSCLC population. The potential impact of the treatments for non-oncology indications for which NICE appraisals were terminated for reasons not related to clinical trial failure on QoL was an incremental improvement of 13% (weighted average). Treatments not available to patients in the UK due to terminated NICE submissions could potentially also reduce the need for rescue therapy, surgery, hospitalization, and transfusions (29, 33, 34, 38), therefore reducing the burden on the NHS, or have a minor positive effect on caregivers’ QoL and their ability to achieve major life goals related to education, work, or family life (27).

The increase in terminated NICE appraisals for multi-indication products limits the availability of innovative healthcare technologies to NHS patients and creates disparity with other European countries, as well as within the UK, where these treatments have been reimbursed and may be available as the SoC. The 23 treatments and indications with terminated NICE appraisals included in the current, UK-based study were analysed across seven other European countries using HTA evaluation reports from national HTA agencies. Up to 21 were accepted for reimbursement in other countries (Figure 2) (62).

**Figure 2:**
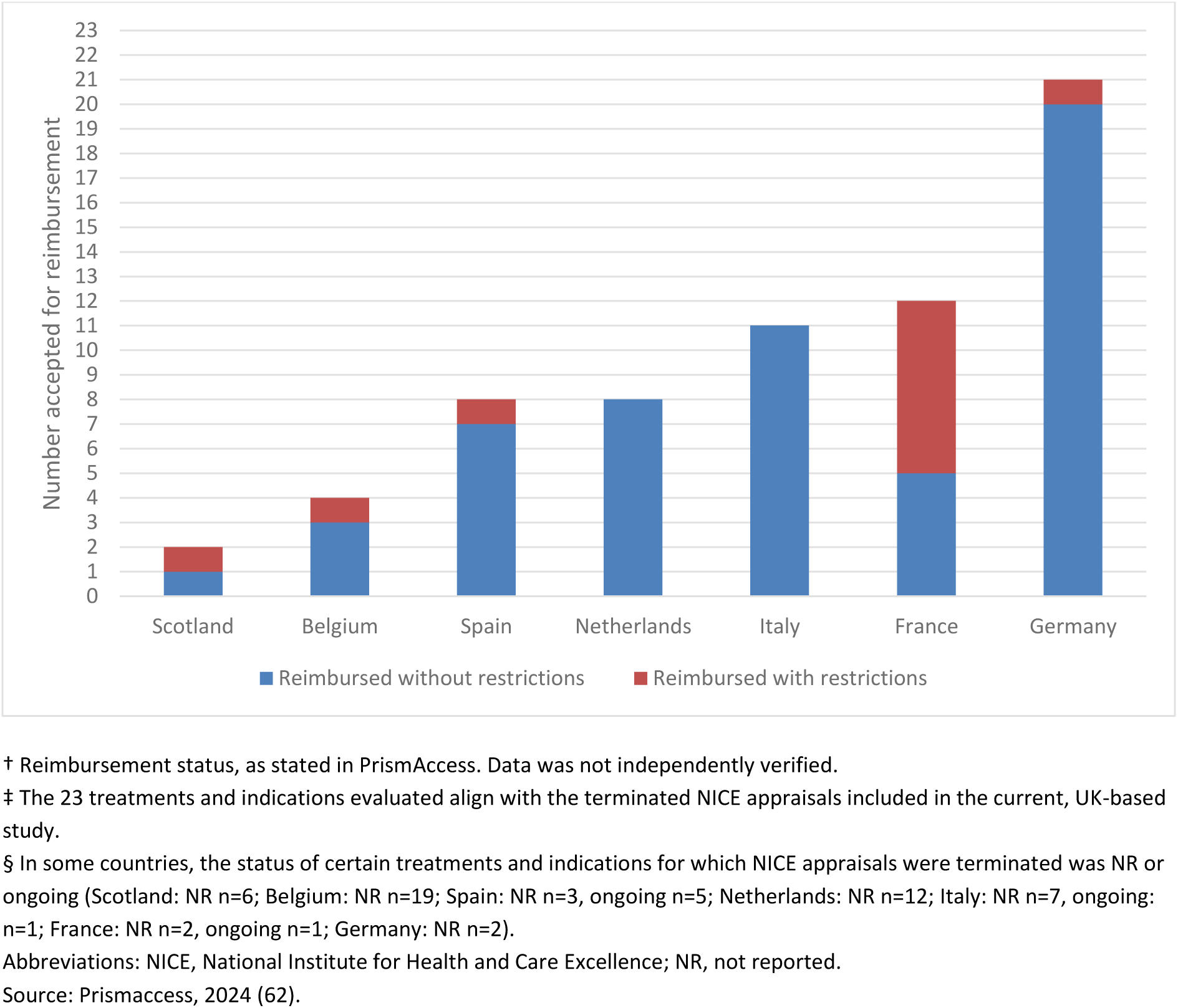
Reimbursement status^†^ of the 23 treatments and indications for which NICE appraisals^‡^ were terminated across eight other European countries^§^

Overall, the European Federation of Pharmaceutical Industries and Associations Patients (EFPIA) Waiting to Access Innovative Therapies (WAIT) Indicator 2023 Survey reported that England is ranked ninth in Europe for total availability of approved medicine (2019–2022), falling behind all other EU5 countries (63). On average, 43% of innovative medicines with marketing authorisation via the centralised European Medicines Agency (EMA) process between 2019 and 2022 (n=72/167 products) were available to patients in the EU, and 40% of available products had limited availability (n=29/72) (63). In England, whilst 56% of products (n=93/167) were available, 49% of available products had limited availability (n=46/93) (63). In comparison, 88% of products (n=147/167) were available in Germany, the country ranked first in Europe for total availability of approved medicine, only 1% of which had limited availability (n=1/147) (63).

The termination of NICE appraisals for multi-indication products for reasons not related to clinical trial failure has prevented NHS patients from accessing innovative healthcare technologies that could provide clinical benefit and are available in other European countries. An example of this is the treatment of adults with chronic rhinosinusitis with nasal polyposis (CRSwNP), a type 2 inflammatory disease of the paranasal mucosa that is associated with significant morbidity and a high symptom burden, such as rhinorrhea, loss of smell, and nasal congestion, which reduces physical and mental HRQoL, including sleep quality (64, 65). Currently available interventions for patients with CRSwNP in the UK are associated with recurrence of nasal polyps and accompanying symptoms, along with a risk of adverse effects, particularly with long-term or repeated use (64), highlighting a clear unmet need for further treatment options. Three different biologics (dupilumab, omalizumab and mepolizumab), all multi-indication products, are licensed in Europe for the treatment of CRSwNP that is uncontrolled with intranasal corticosteroids (66–68). All have demonstrated significant improvements in the reduction of nasal polyps and accompanying symptoms (65, 69–71). The use of biologics is recommended by British and international guidelines for a clearly defined group of patients (72, 73), but none are available on the NHS due to terminated NICE appraisals (74–76). This means that patients in the UK have no access to any biologic therapy in this indication, despite these treatments being widely reimbursed outside the UK; for example, dupilumab is currently reimbursed for the treatment of patients with CRSwNP in 30 different countries, including all other EU5 countries (France, Germany, Italy and Spain), Canada, the US, and Japan.

The impact of the increase in terminated NICE appraisals for products with multiple indications for reasons not related to clinical trial failure on OS and PFS in oncology may contribute to the poor global ranking of the UK for cancer survival; the 2024 analysis of international data by the Less Survivable Cancers Taskforce concluded that the UK ranks 28^th^, 26^th^ and 21^st^ for five-year survival of lung cancer, pancreatic cancer, and liver cancer, respectively, out of 33 countries of comparable wealth and income levels (77). Of the eight terminated NICE appraisals for oncology indications assessed by this study, three were for patients with lung cancer (TA436 (15, 16)), pancreatic cancer (TA750 (21)) or liver cancer (TA609 (22)). The termination of NICE appraisals for these indications means that patients in the UK are unable to access innovative treatments which may have impacted their survival.

In countries such as France and Germany, the value of treatments is assessed primarily on the clinical benefits to patients, with pricing processes and negotiations conducted separately (78, 79). In countries such as Italy and Spain, multiple elements of value are assessed, including both clinical and cost-effectiveness (78, 79). However, in the UK, the incremental clinical benefits and costs are combined into one measure, the incremental cost-effectiveness ratio, and value is assessed based on cost-effectiveness analysis (79, 80), with access decisions typically based on a willingness-to-pay threshold (80), which may hinder the reimbursement of products with multiple indications. The UK uniform pricing policy limits recognition of the value of multi-indication products, as the lowest cost-effective price determined for any indication must be implemented across all indications (5). Therefore, the UK uniform pricing policy is likely to be a factor for manufacturer-driven termination of appraisals to NICE. Given the approach to value assessment and pricing in the UK, more flexible pricing options are needed to ensure that access to treatments is in line with other countries in Europe.

A potential solution is IBP agreements, which can enable price to vary by indication and reflect the therapeutic value of the treatment (6). In a 2020 global survey across 16 countries (N=73; respondents represented industry [37%], payers [27%], regulators [16%] and academics [10%], among other stakeholders), including the UK (n=17), 78% of respondents agreed that some form of IBP agreements would be a good thing (81). More than half of all respondents (57%) thought that all stakeholders could stand to gain from IBP agreements (81). If successfully applied, IBP agreements can contribute towards better resource allocation, improve patient access to treatments, and incentivise research and development (6, 82), which could increase price competition at the indication-level, ultimately resulting in lower prices and better value to health systems (6). IBP agreements could take the form of a single price based on a weighted average of value and usage across indications (6). Alternatively, discount levels or rebates that vary by indication could be applied, or agreements between manufacturers and payers that adjust price according to realised performance (6). For example, value-based discounts have applied by individual insurers for dupilumab in the United States, Germany and Australia (83), the latter of which assesses value based on cost-effectiveness analysis like the UK. Adoption of IBP agreements could help to minimise the termination of NICE appraisals for products with multiple indications by allowing price to remain commensurate with the therapeutic value of the product in each indication, an increasingly urgent issue as the number of products with multiple indications rises, with 46.3% of NICE appraisals in development for multi-indication products as of September 2023 (84).

This study has several limitations. The results presented reflect potential QALY and survival foregone based on targeted literature searches conducted to identify data on the health impact of the interventions, as there are no manufacturer evidence submissions for the identified terminated appraisals. Due to the large number of indications, a targeted literature search was performed for each indication rather than a systematic literature review, which may have resulted in some relevant studies being missed in the analysis. Several assumptions were made due to lack of data: in the absence of direct evidence, studies published in a similar indication were used as proxies; an exponential distribution was employed for all survival analyses; and when calculating the annual incremental QALY, an even distribution across the modelled time horizon was assumed. Therefore, accurate estimates of QALY and survival loss are very difficult to assess. The intent behind this analysis is to provide a broad estimate of the health impact of the lack of access to treatments with multiple indications for which NICE appraisals have been terminated on the UK population, to stimulate discussion about the UK pricing environment.

## Conclusion

The increasing number of NICE terminations, particularly for multi-indication products, means that patients are unable to access therapies that could potentially lengthen their lives and increase their QoL. Addressing access barriers to bring access in line with other European countries, through solutions like IBP agreements, could provide an opportunity to improve UK public health.

## Acknowledgements

This study was funded by Sanofi. The authors received writing support from Eleanor Ward and Sophie Doran, as well as quality checking/validation support for methods and calculations from Margaux Cornell and Carina Bektur, employees of Clarivate. The authors also received data extraction and manuscript review support from Adele Schulz.

## Author Declarations

The authors declare that all relevant ethical guidelines have been followed, all necessary IRB and/or ethics committee approvals have been obtained, all necessary patient/participant consent has been obtained and the appropriate institutional forms archived. The authors declare their understanding that any clinical trials described should be registered with an internationally recognized trial registry and the trial ID included in the manuscript.

## Competing Interest Statement

The study was funded by Sanofi, and Helen Mitchell, Clement Halin, and Richard Hudson are employees and stockholders of Sanofi. Qian Xin, Jack Hide, Swarali Sunil Tadwalkar, and Sabera Hashim are employees of Clarivate, an institution which received funding from Sanofi to conduct the analysis and for writing support. The authors indicated no further potential conflicts of interest.

## Funding Statement

This study was funded by Sanofi, with funding used to conduct the analysis and for writing support from Clarivate.

## Data Availability

All data produced in the present study are available upon reasonable request to the authors.

## Notes

### Competing Interest Statement

All authors have completed the ICMJE uniform disclosure form at www.icmje.org/coi_disclosure.pdf and declare: this work was funded by Sanofi with funding used to conduct the analysis and for writing support from Clarivate; HM, CH and RH are all employees and stock holders of Sanofi; authors have no other relationships or activities that could appear to have influenced the submitted work.

