## Appendix 1 for "Is the United Kingdom (UK) medicines pricing policy failing patients? The impact of terminated National Institute for Health and Care Excellence (NICE) appraisals for multi-indication products on patients"

*Table 8: Studies included in the final analyses*

| Reference number | Study | Type of analysis | Study design | Country/region | Indication | Treatment | Comparator | Notes |
|---|---|---|---|---|---|---|---|---|
| **TA436** | Li et al, 2021 (1) | QALY | CEA | Japan | NSCLC | Bevacizumab | Erlotinib | |
| | Li et al, 2023 (2) | Survival | Meta-analysis | International | NSCLC | Bevacizumab | Erlotinib | |
| **TA353** | Hinde et al, 2016 (3) | QALY & survival | CEA | UK | Ovarian cancer | Bevacizumab | Chemotherapy | |
| | Chappell et al, 2016 (4) | Survival | CEA | International | Ovarian cancer | Bevacizumab | Chemotherapy | |
| **TA717** | Chen et al, 2023 (5) | QALY | CEA | China | Follicular lymphoma | Duvelisib | Bendamustine plus rituximab | |
| | Patel et al, 2019 (6) | Survival | Literature review | International | Follicular lymphoma | Duvelisib | Ofatumomab | |
| **TA750** | Mirzayeh Fashami et al, 2023 (7) | QALY & survival | CEA | Canada | Pancreatic cancer | Olaparib | Placebo | |
| **TA609** | Zheng et al, 2020 (8) | QALY & survival | CEA | US | Hepatocellular cancer | Ramucirumab | Placebo | |
| **TA933** | Qi et al, 2021 (9) | QALY | CEA | US | Large B-cell lymphoma | Tisagenlecleucel | Salvage chemotherapy | |
| | Maziarz et al, 2022 (10) | Survival | ITC | International | Large B-cell lymphoma | Tisagenlecleucel | Historical treatments | |
| **TA901** | Liu et al, 2023 (11) | QALY & survival | CEA | US | Metastatic or recurrent cervical cancer | Cemiplimab | Chemotherapy | |
| **TA648** | Yong et al, 2021 (12) | QALY | CEA | US | Chronic rhinosinusitis | Dupilumab | Aspirin desensitization therapy | |
| | Oykhman et al, 2022 (13) | QoL | Meta-analysis | International | Chronic rhinosinusitis | Dupilumab | Endoscopic sinus surgery | |
| **TA636** | ICER, 2021 (14) | QALY | CEA | International | Myasthenia gravis | Eculizumab | Placebo | |
| | Muppidi et al, 2019 (15) | Other† | RCT | International | Myasthenia gravis | Eculizumab | Placebo | |
| **TA647** | Pittock et al, 2019 (16) | QoL | RCT | International | Neuromyelitis optica | Eculizumab | Placebo | |
| | Aungsumart et al, 2020 (17) | QALY | CEA | Thailand | Neuromyelitis optica | Rituximab | Azathioprine | No direct evidence was found for eculizumab in the treatment of |

| | | | | | | | | |
|---|---|---|---|---|---|---|---|---|
| | | | | | | | | neuromyelitis optica, so data on rituximab in the treatment of neuromyelitis optica were used as a proxy |
| **TA843** | Cappellini et al, 2020 (18) | Other† | RCT | International | Anaemia | Luspatercept | Placebo | |
| **TA844** | Olivia et al, 2021 (19) | QoL | RCT | International/US | Anaemia | Luspatercept | Placebo + BSC | |
| | Htut et al, 2023 (20) | Other† | Meta-analysis | International | Anaemia | Luspatercept | Placebo/erythropoietin stimulating agent | |
| **TA845** | ICER, 2016 (21) | QALY | CEA | US | Severe asthma with eosinophilia | Mepolizumab | SoC | No direct evidence was found for mepolizumab in the treatment of EGPA, so data on mepolizumab in the treatment of severe asthma with eosinophilia were used as a proxy |
| **TA846** | Von Maltzahn et al, 2021 (22) | QoL | RCT | International | HES | Mepolizumab | Placebo | |
| | ICER, 2016 (21) | QALY | CEA | USA | Severe asthma with eosinophilia | Mepolizumab | SoC | No direct evidence was found for mepolizumab in the treatment of HES, so data on mepolizumab in the treatment of severe |

| | | | | | | | | |
|---|---|---|---|---|---|---|---|---|
| | | | | | | | | asthma with eosinophilia were used as a proxy |
| **TA637** | Ross et al, 2016 (23) | QALY | CEA | US | Diabetic retinopathy | Ranibizumab | Bevacizumab | |
| | Vijayan et al, 2017 (24) | QoL | Observational study | India | Diabetic retinopathy | Ranibizumab | Intravitreal bevacizumab therapy + focal laser | |
| **TA840** | Ong et al, 2023 (25) | QALY | CEA | Singapore | Chronic GvHD refractory to corticosteroids | Ruxolitinib | Best alternative therapy | |
| **TA839** | Ong et al, 2023 (26) | QALY | CEA | Singapore | Acute GvHD in Patients - 12 Years of Age | Ruxolitinib | Best alternative therapy | |
| **TA826** | Petryszyn et al, 2020 (27) | QALY | CEA | Poland | Ulcerative colitis | Vendolizumab | SoC | |
| | NICE, 2014 (28) | QoL | NICE TA | UK | Ulcerative colitis | Vendolizumab | Placebo | |
| **TA940** | Vu et al, 2022 (29) | QoL | RCT | USA | Generalized myasthenia gravis | Ravulizumab | Placebo | |
| | Tice et al, 2022 (30) | QALY | CEA | International | Myasthenia gravis | Eculizumab | Placebo | |
| **TA941** | Clardy et al, 2024 (31) | Other† | Meta-analysis | International | Neuromyelitis optica | Ravulizumab | Eculizumab, satralizumab or inebilizumab | |
| | Aungsumart et al, 2020 (17) | QALY | CEA | Thailand | Neuromyelitis optica | Rituximab | Azathioprine | |
| **TA938** | Sanofi data on file, 2024 (32) | QALY | CEA | UK | EGPA | Dupilumab | SoC | |
| **TA899** | Eric et al, 2020 (33) | QALY | CEA | US | Treatment-resistant depression | Esketamine | Oral antidepressents | |
| | Jamieson et al, 2023 (34) | QoL | Meta-analysis | International | MDD with active suicidal ideation and intent | Esketamine + SoC | Placebo+ SoC | |
| **TA910** | Mital et al, 2023 (35) | QALY | CEA | US | Adolescents with | Semaglutide | No treatment | |

| | | | | | |
|---|---|---|---|---|---|
| | | | | | severe obesity |

† Additional qualitative analysis was conducted to explore other potential health impacts that could not be quantified at the population level due to a lack of data.

Abbreviations: BSC, best supportive care; CEA, cost-effectiveness analysis; EGPA, eosinophilic granulomatosis with polyangiitis; GvHD, graft vs. host disease; HES, hypereosinophilic syndrome; ICER, Institute for Clinical and Economic Review; ITC, indirect treatment comparison; MDD, major depressive disorder; NICE, National Institute for Health and Care Excellence; NR, not reported; QALY, quality-adjusted life year; QoL, quality of life; RCT, randomized control trial; Soc, standard of care; TA, technology assessment; UK, United Kingdom; US, United States.
